# Towards robust clinical genome interpretation: developing a consistent terminology to characterize disease-gene relationships - allelic requirement, inheritance modes and disease mechanisms

**DOI:** 10.1101/2023.03.30.23287948

**Authors:** Angharad M Roberts, Marina T. DiStefano, Erin Rooney Riggs, Katherine S Josephs, Fowzan S Alkuraya, Joanna Amberger, Mutaz Amin, Jonathan S. Berg, Fiona Cunningham, Karen Eilbeck, Helen V. Firth, Julia Foreman, Ada Hamosh, Eleanor Hay, Sarah Leigh, Christa L. Martin, Ellen M. McDonagh, Daniel Perrett, Erin M. Ramos, Peter N. Robinson, Ana Rath, David van Sant, Zornitza Stark, Nicola Whiffin, Heidi L. Rehm, James S. Ware, the Gene Curation Coalition

## Abstract

**PURPOSE:** The terminology used for gene-disease curation and variant annotation to describe inheritance, allelic requirement, and both sequence and functional consequences of a variant is currently not standardized. There is considerable discrepancy in the literature and across clinical variant reporting in the derivation and application of terms. Here we standardize the terminology for the characterization of disease-gene relationships to facilitate harmonized global curation, and to support variant classification within the ACMG/AMP framework.

**METHODS:** Terminology for inheritance, allelic requirement, and both structural and functional consequences of a variant used by Gene Curation Coalition (GenCC) members and partner organizations was collated and reviewed. Harmonized terminology with definitions and use examples was created, reviewed, and validated.

**RESULTS:** We present a standardized terminology to describe gene-disease relationships, and to support variant annotation. We demonstrate application of the terminology for classification of variation in the ACMG SF 2.0 genes recommended for reporting of secondary findings. Consensus terms were agreed and formalized in both sequence ontology (SO) and human phenotype ontology (HPO) ontologies. GenCC member groups intend to use or map to these terms in their respective resources.

**CONCLUSION:** The terminology standardization presented here will improve harmonization, facilitate the pooling of curation datasets across international curation efforts and, in turn, improve consistency in variant classification and genetic test interpretation.

## Introduction

The clinical application of genomic data is reliant upon a robust understanding of the relationships between locus, genotype, mechanism, and disease phenotypes ^1^.

Assessment of the evidence that variants in a gene cause a particular monogenic disease is critical for variant classification, particularly for clinical application ^2-4^. The American College of Medical Genetics and Genomics and the Association for Molecular Pathology (ACMG and AMP) have issued standards and guidelines for the clinical interpretation of sequence variants which have now been widely adopted internationally ^2^. These standards and further subsequent guidance make clear that the first step in the classification and interpretation of a variant is the robust assessment of disease-gene validity ^4^; without a clear understanding of the gene’s role in disease, variant assessment criteria cannot be accurately applied ^5,6^. Using an incorrectly classified variant for family cascade testing and the delivery of screening, treatment, or reproductive choices can have severe adverse consequences.

Historically, disease genes were identified by linkage studies in large families ^7^ often using polymorphic markers in close proximity to the gene responsible for the disease. Subsequently candidate gene studies based on known or hypothesized disease mechanisms became commonplace ^8^. Until as recently as 10 years ago it was not fully appreciated that individually-rare genetic variants are collectively extremely common, and many studies did not adequately control for background genetic variation in the population. Inadequately small control cohorts, often just 100 chromosomes, were used for assessment of novel disease genes and variants, and consequently both the literature and disease databases were flooded with assertions of gene-disease relationships and variant pathogenicity that have not proven robust over time ^9-11^. Recent years have seen concerted efforts to correct this bias, employing large, publicly available population databases such as ExAC, and later gnomAD ^12,13^, as control cohorts, and applying standardized approaches to reinterpret evidence for gene-disease relationships ^5^ and variant pathogenicity ^2^

Many groups are invested in curation of disease-gene validity, including academic and healthcare centers, private companies, and consortia. The Gene Curation Coalition (GenCC) is a coalition aiming to harmonize approaches amongst these entities to ensure gene-level curated resources are comparable and interoperable, and to provide access to structured representations of consensus data. As a first undertaking the GenCC developed a consensus term set for grading gene–disease validity, and developed a unified database to display curated gene–disease validity assertions from its members (the Clinical Genome Resource (ClinGen), DECIPHER, Gene2Phenotype (G2P), Transforming Genetic Medicine Initiative (TGMI), MedlinePlus Genetics, Genomics England PanelApp (PanelApp), PanelApp Australia, Online Mendelian Inheritance in Man (OMIM), Orphanet, Ambry Genetic, Illumina, Invitae, Mass General Brigham Laboratory for Molecular Medicine (LMM), Myriad Women’s Health, HUGO Gene Nomenclature Committee (HGNC), Franklin by Genoox, King Faisal Specialist Hospital and Research Center, and PharmGKB). This database can be likened to a “ClinVar for genes” in that members can submit assertions of disease association but in this case for genes, not variants. The GenCC database provides a single route of access to comprehensive aggregated assertions (https://search.thegencc.org) ^14^ and currently contains over 16,911 gene–disease assertions on 4704 unique genes from 12 submitters. OMIM will connect its large dataset to the GenCC in real time via a submission API being launched soon. Resolution of gene-disease validity discrepancy across GenCC submitters is ongoing using a manual review process.

As the next step to facilitate harmonized gene-disease validity assessments and support variant classification within the ACMG/AMP framework, GenCC has focused on developing standardized terminology for the characterization of disease mode of inheritance, allelic requirement, and disease-associated variant consequences. Currently, groups utilize different terminology to describe these three characteristics. The considerable discrepancy in the derivation and application of these terms generates confusion and risks discordant assertions about pathogenicity of different classes of variants.

Although there is a close conceptual relationship between inheritance and allelic requirement, they are distinct and serve different purposes. Inheritance is used for describing the mode of transmission of a phenotype e.g. autosomal dominant, and is particularly applicable in the clinical setting for communicating recurrence risk, and to guide family screening and reproductive advice. Allelic requirement describes how many alleles must be impacted to cause the relevant disease e.g. one allele (monoallelic) in dominant disease, and both alleles (biallelic) in recessive disease. It is necessary for variant annotation pipelines and to determine if a given variant in a specific context is relevant to the phenotype of the patient e.g. a single heterozygous variant (in the absence of compound heterozygosity) may provide a diagnosis for dominant disease, but is insufficient to explain recessive disease, where a second contributory variant or alternative cause should be sought.

Disease-associated variant consequence in particular can be useful when evaluating novel variants in validated disease-associated genes. Is the predicted consequence of the novel variant consistent with that of previously reported disease-causing variants or with the mechanism of disease (if known)? For many genes the mechanism of action will not yet be known even if the gene-disease validity has been confirmed. Understanding the consequence of known pathogenic variants is a useful intermediate step that can aid variant classification and inform understanding of the mechanism of disease. For example, the consequence of a nonsense-mediated-decay (NMD) competent nonsense variant is a reduction in the amount of gene product produced If all known pathogenic variants in a gene are nonsense, not only can we predict that a novel NMD competent nonsense variant identified in a patient would have a high likelihood of being pathogenic, we can postulate that any novel variant with the same consequence (reduction in the amount of gene product produced), would also be pathogenic. Considering a NMD competent nonsense variant, a whole gene deletion or other variants resulting in premature termination codons (PTCs) including frameshift and essential splice site variants, could be considered equivalent, assuming they are located in required exons and sufficiently upstream to lead to lead to the same effect. Regulatory variants in non-coding regions that abolish protein expression can also have equivalent downstream effects^15^. An advantage of using disease-associated variant consequence in an era of increasing appreciation of the clinical importance of non-coding variants is that it is applicable across both coding and non-coding region variants.

Having structured data representations compatible across platforms to describe inheritance, allelic requirement, and disease-associated variant consequence can help avoid duplication of effort, facilitate manual annotation, and can be more readily incorporated into automated analysis pipelines.

Many groups provide resources to disseminate gene-disease curations, with varying levels of detail - e.g. some assess whether there is a gene-disease relationship, while others add details of disease mechanisms. Each group may use different terminology to describe inheritance (e.g. autosomal dominant, autosomal recessive, etc.), allelic requirement, and both structural and functional consequences of a variant ^16-19^.

The Gene Curation Coalition (GenCC) promotes use of standardized terms for structured representation of gene-disease relationships, including strength of evidence for the gene-disease association, disease mode of inheritance, allelic requirement, structural and functional consequences of genetic variation, and mechanism of pathogenicity. This will allow for harmonized terminology across genetic resources, and aid variant curation, classification, and reporting.

## Methods

### Consensus development panel

The process followed for developing and testing the terminology is outlined in figure 1.

**Fig 1.**
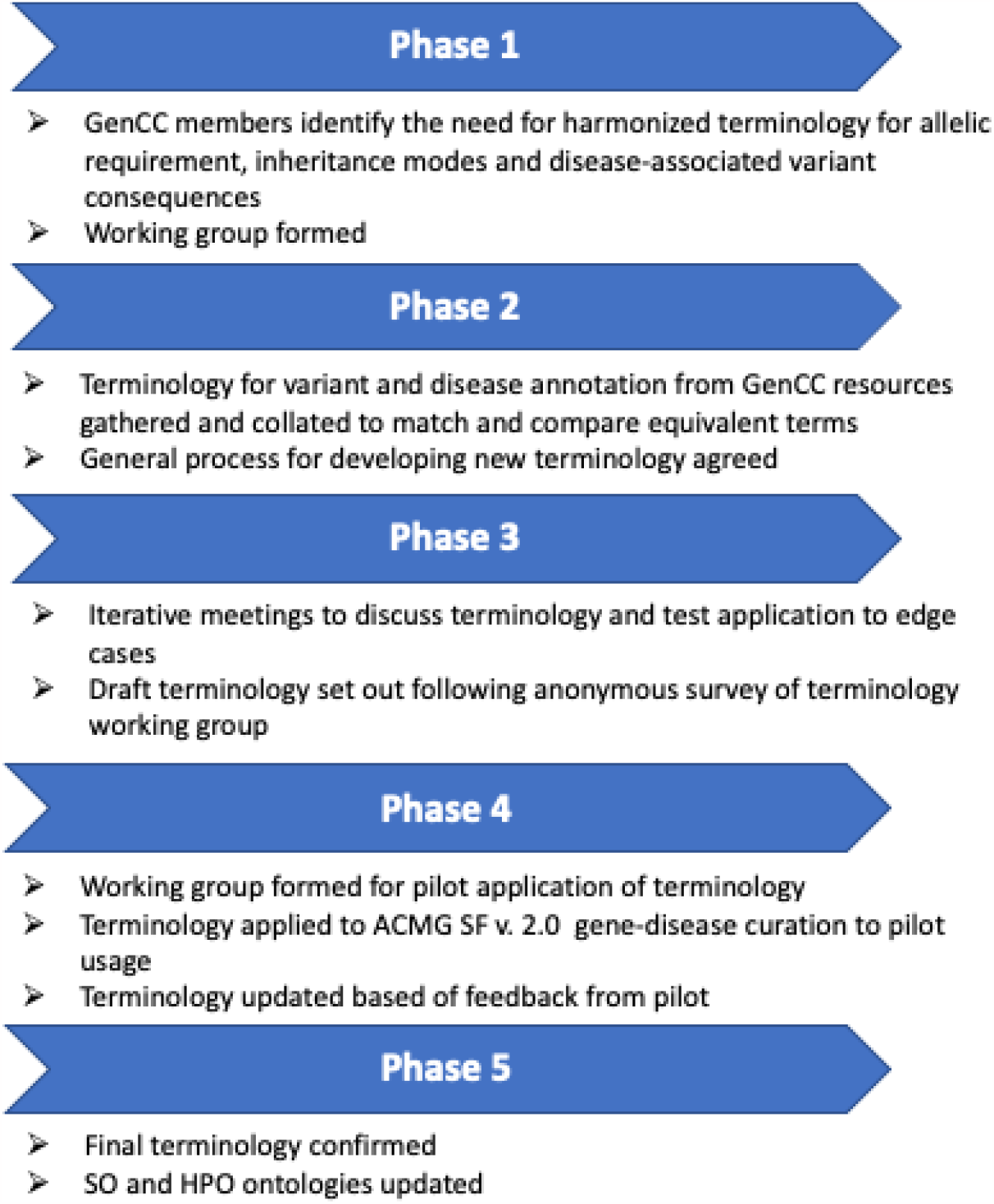
Process for developing and testing the terminology

The GenCC includes experts in the identification and evaluation of variants from diverse settings, including clinical and research contexts, academic and commercial laboratories, software and resource developers, and organizations maintaining current nomenclature standards. An initial meeting and scoping exercise identified the need for a harmonized framework and terminology for inheritance, allelic requirement, and disease-associated variant consequences. The ultimate goal will be to understand precise mechanisms of disease and predict precise functional consequences of variants, but we have not yet developed a structured ontology for mechanism given the enormous diversity of possible functional effects of genetic variation. The groupings of predictable gene product changes will allow for consistent variant prioritization pending further functional characterization. The panel met by monthly conference call between February 2019 and September 2022. Existing terminology used by GenCC members and partners was reviewed and collated (supp table 1 & 2). Existing terms for allelic requirement and inheritance coalesce around Human Phenotype Ontology (HPO) terms^20^, while terms for disease-associated variant classification coalesce around Sequence Ontology (SO) terms^21^.

An updated framework and ontology were developed through iterative discussion and survey. All members of the panel reviewed and approved the final terminologies. Consensus was defined as agreement among most (>80%) members of the panel. The penultimate draft of the ontology and framework was generated following an anonymous online survey of the panel. Changes were made as considered appropriate based on feedback from the pilot working group.

### Pilot curation working group

A working group of clinical geneticists with experience in the identification and evaluation of variants in both the clinical and research settings was formed to pilot curation of the 59 genes (66 disease-gene pairs) included in the ACMG Recommendations for Reporting of Incidental Findings in Clinical Exome and Genome Sequencing version 2.0 ^22^(current at the time of study) using the new terminology and variant consequence matrix. The template supplied to curators in the pilot working group and the final outputs are available in the supplemental materials.

## Results

### Terminology for inheritance and allelic requirement

Most coalition members used separate inheritance and allelic requirement terms, with the exception of G2P, Genomics England PanelApp and PanelApp Australia which use only allelic requirement terms.

### Inheritance

Review of existing inheritance terms identified substantial consistency in high level terms used, e.g. autosomal recessive/autosomal dominant, with the exception of G2P, Genomics England PanelApp and PanelApp Australia which use monoallelic/biallelic as stem terms for both inheritance and allelic requirement (See supplementary table 1). Some groups also use modifier terms which can be applied to some or all stem terms to add further granularity, for example indicating whether a gene is imprinted, the penetrance of variation in a gene, or whether pathogenic variants in a given gene are typically *de novo* or mosaic. There was less consistency between GenCC members in whether modifiers were used and in modifier terms themselves. Most consortium members’ existing stem terminology mapped broadly to HPO inheritance terms. It was also noted that there were also some redundant HPO inheritance terms.

It was agreed to collaborate with HPO to update HPO inheritance terms (children of HP:0000005), and to harmonize on the first level. HPO inheritance terms are a relatively small ontology intended to describe the mode of inheritance, and contain terms such as ‘Autosomal dominant inheritance HP:0000006’. Original HPO inheritance terms also included several terms that were not true modes of inheritance, but did provide useful genetic information, such as ‘Genetic anticipation HP:0003743’. Three new subcategories of Mode of Inheritance HP:0000005 were created (Table 1a). Mendelian Inheritance (HP:0034345) retains only true inheritance terms (Table 1b), while Inheritance Modifier (HP:0034335) was coined to record descriptors and optional specifications such as ‘Displays anticipation’ or ‘Typically *de novo’* that provide relevant, useful information and can be used in conjunction with any inheritance term (Table 1c), and Complex inheritance HP:0001426 captures complex disease due to mixture of non-Mendelian genetic determinants possibly together with environmental factors. HPO Mendelian inheritance terms were extended to capture all required information for our purposes, for example adding new terms to describe genes encoded in pseudoautosomal regions (PAR recessive HP:0034341, PAR dominant HP:0034340). Following rigorous review, iterative discussion and survey, a final list of inheritance terms and modifiers was agreed upon (Table 1a&b).

**Table 1a.**
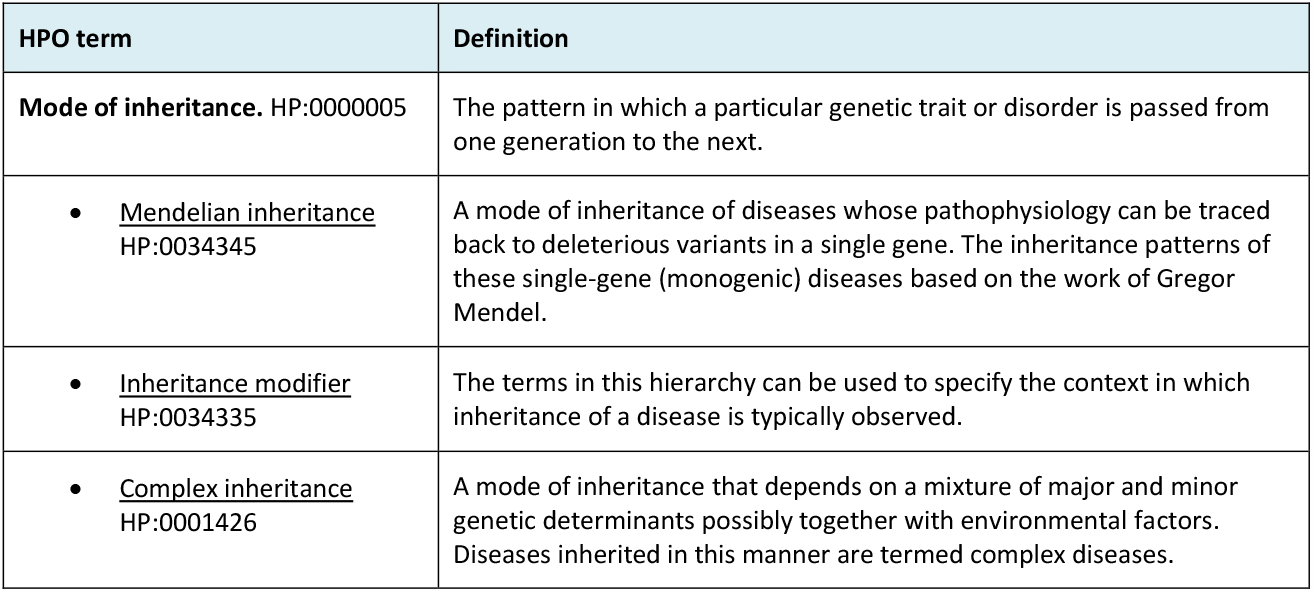
New hierarchy of HPO mode of inheritance child terms and definitions. Abbreviations: HPO - Human phenotype ontology

| HPO term | Definition |
| --- | --- |
| **Mode of inheritance.** HP:0000005 | The pattern in which a particular genetic trait or disorder is passed from one generation to the next. |
| <u>Mendelian inheritance</u><br>HP:0034345 | A mode of inheritance of diseases whose pathophysiology can be traced back to deleterious variants in a single gene. The inheritance patterns of these single-gene (monogenic) diseases based on the work of Gregor Mendel. |
| <u>Inheritance modifier</u><br>HP:0034335 | The terms in this hierarchy can be used to specify the context in which inheritance of a disease is typically observed. |
| <u>Complex inheritance</u><br>HP:0001426 | A mode of inheritance that depends on a mixture of major and minor genetic determinants possibly together with environmental factors. Diseases inherited in this manner are termed complex diseases. |

**Table 1b.**
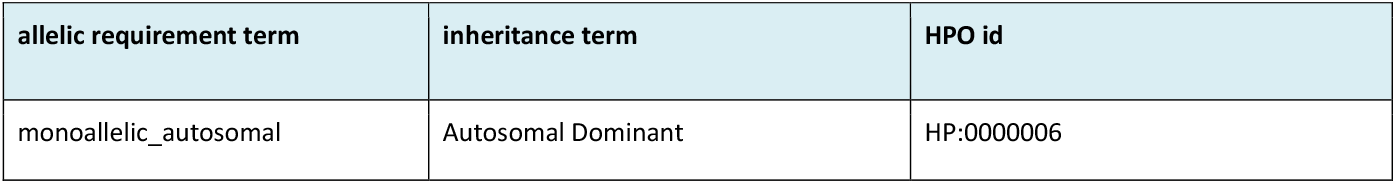
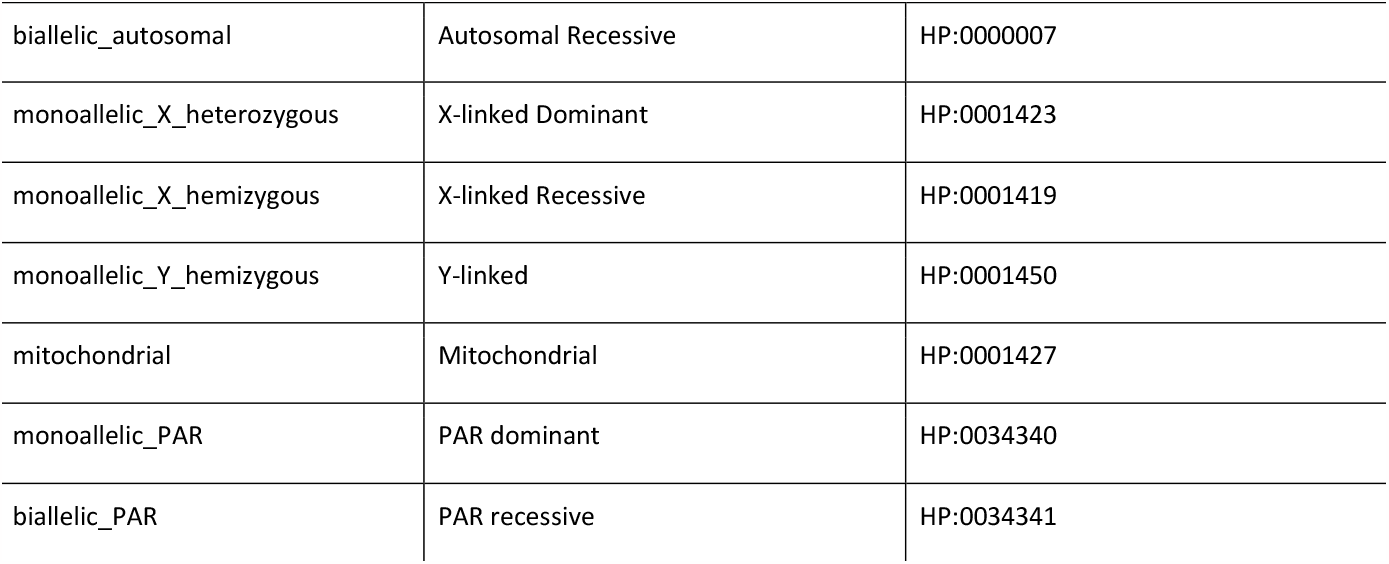
Harmonized allelic requirement and Mendelian inheritance terms, child terms of HP:0034345. Abbreviations: HPO - Human phenotype ontology, PAR - pseudoautosomal region.

**Table 1c.**
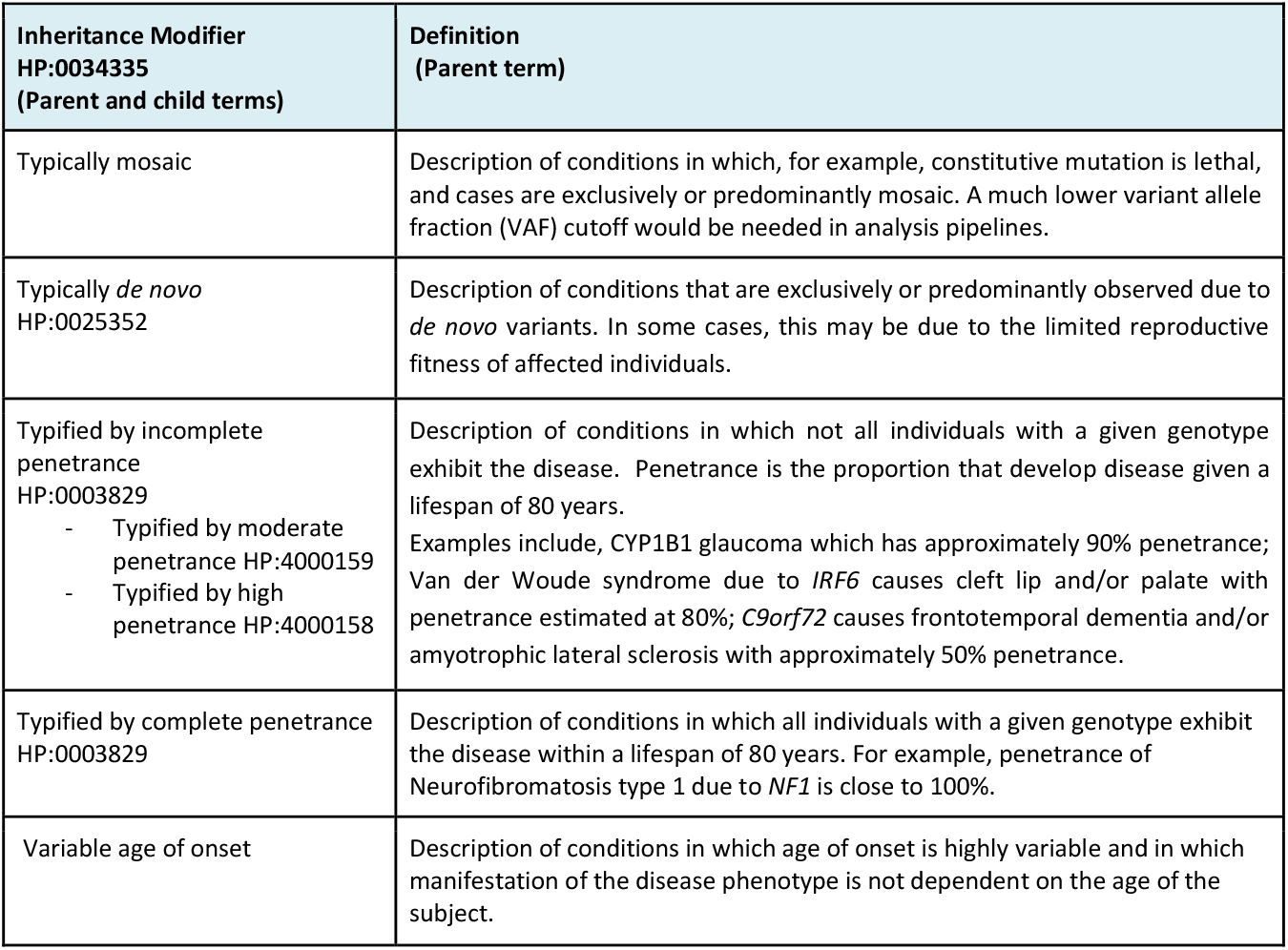
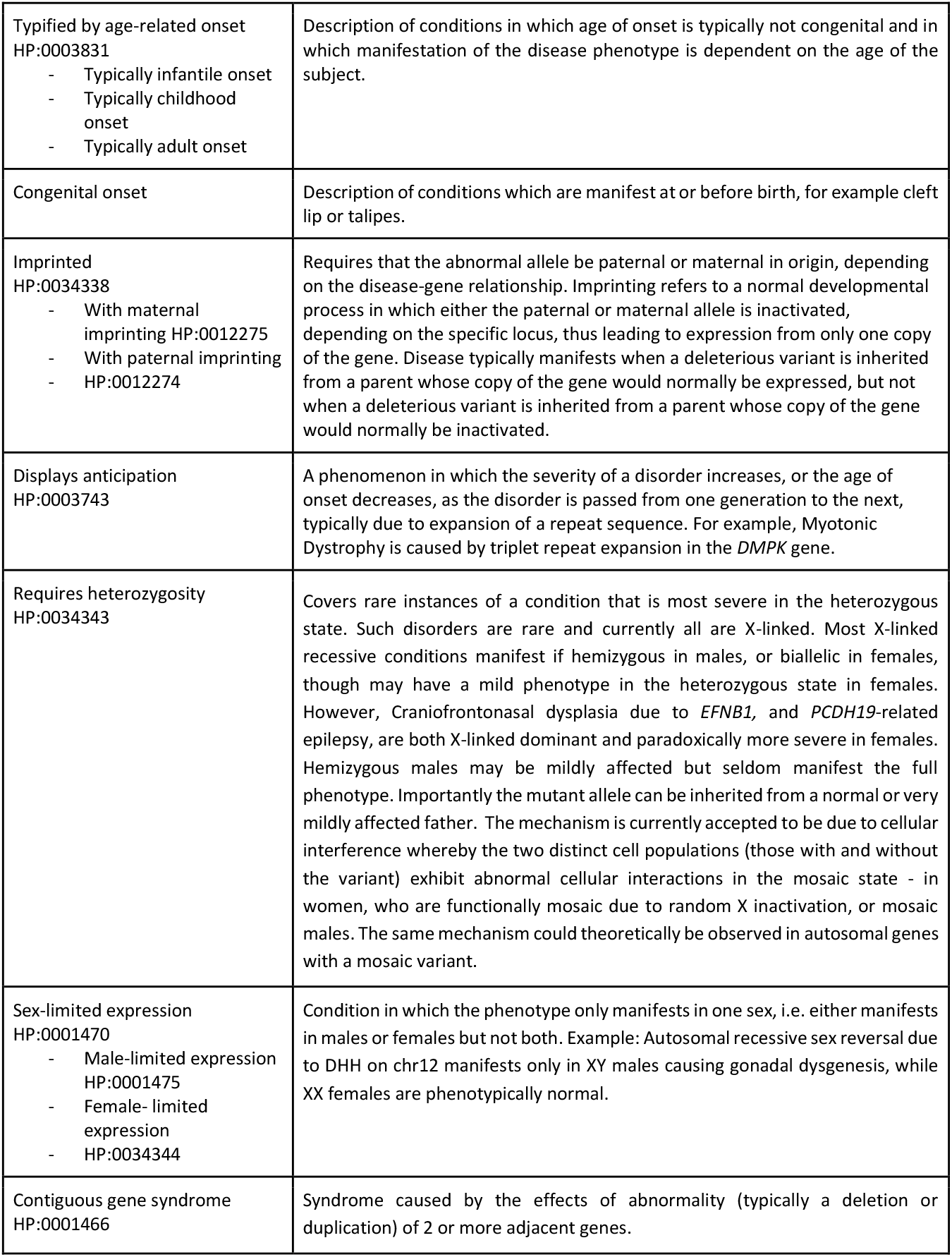
Inheritance modifier terms- these optional terms can be combined with either inheritance terms or allelic requirement terms to provide additional information about the relationship of a disease-gene pair.

### Allelic requirement

Consistency was also identified between GenCC groups in the use of existing allelic requirement terms, with most groups using monoallelic/biallelic as stem terms, with the exception of DECIPHER, which used zygosity terms for variants e.g. ‘heterozygous’ (see Supplementary table S2). Again there was less consistency between members in the use of modifiers, ranging from no modifiers to structured modifier terms to long narrative modifiers. HPO did not previously have terms for allelic requirement, so a proposed terminology was derived from first principles and terms most commonly used among GenCC members. Following iterative discussion and survey, a final list of allelic requirement terms was agreed upon that captures whether the disease results from monoallelic or biallelic variation; whether encoded on an autosome, a sex chromosome, or the pseudoautosomal region; and for monoallelic diseases on the X chromosome whether they manifest when heterozygous, or hemizygous.

### Alignment of inheritance and allelic requirement terms

Finalized terms for inheritance and allelic requirement, which had been derived independently of each other, were aligned and adjusted as needed such that each mode of inheritance had an accompanying allelic requirement term to describe the context necessary to cause disease (table 1). Allelic requirement terms were added to HPO as allelic requirement synonyms of Mendelian inheritance terms and share HPO identifiers with the corresponding inheritance term. Cross-cutting inheritance modifiers generated for use with inheritance terms above were expanded by consideration of edge cases such as Craniofrontonasal dysplasia due to *EFNB1* which requires heterozygosity^23^. Modifier terms were refined to be compatible with all inheritance and allelic requirement terms by iterative discussion, to enable recording of data important to reproductive advice and family screening. Examples to illustrate the applications of these terms are shown in Box 1

#### Box 1

Example applications of harmonized allelic requirement terms

An X-linked dominant condition would be curated as monoallelic_X_heterozygous and we would understand that those diseases are typically observed in females though may less commonly manifest in males when hemizygous (or indeed in females when homozygous/compound heterozygous) though this may be more severe or lethal.

*e*.*g. Rett syndrome due to MECP2, a severe neurodevelopmental disorder that occurs almost exclusively in females* ^*24*^. *Rarely, classically affected males with somatic mosaicism or an extra X chromosome have been described, usually with an earlier onset of symptoms*.

An X-linked recessive condition would be curated as monoallelic_X_hemizygous and we would understand that those diseases are most commonly observed in males and may not fully manifest when heterozygous in females. Though they can manifest with ameliorated phenotype, or manifest if skewed inactivation etc., we intend that this is implicit in the term as characteristic of many sex-linked disorders, and do not anticipate that an additional modifier term is needed to communicate this, unless the heterozygous phenotype is sufficiently distinct as to be classified as a different disease entity. It is also implicit that monoallelic variants in females with chromosomal anomalies (e.g. 45,X) and biallelic variants in females would also meet the allelic requirement.

*e*.*g*. Duchenne muscular dystrophy (DMD)^25^. Males are affected by childhood onset skeletal muscular dystrophy and later onset cardiomyopathy, typically in adolescence. Approximately 10% of female carriers have some (typically mild) symptoms and 20% have cardiac involvement on investigation. Carrier females and females with biallelic variants with the full DMD phenotype have been reported.

Terms are specific to each disease-gene pair. Considering a hypothetical example of a gene on the X-chromosome in which biallelic or hemizygous monoallelic variation causes congenital structural heart abnormalities, but a heterozygous monoallelic variant typically presents with late onset cardiomyopathy, this might be coded as monoallelic_X_hemizygous for congenital heart disease, and appropriate filtering applied in a developmental disorders panel for diagnosis of an infant, and monoallelic_X_heterozygous (age-related onset) for cardiomyopathy, with different variant filtering applied for a cardiac gene panel analysis in an adult. This scenario is an example in which genetic variation in a single gene can give rise to separate disease phenotypes that are curated as distinct entities and has the advantage of tracing the evidence for each disease association.

### Variant classes and disease-associated variant consequences

Structured terminology to capture disease associated variant classes (e.g. missense, nonsense, etc.) is required to support variant classification within the ACMG/AMP framework, both to infer which variant classes are likely to be relevant, and to support assessment of relevance of functional evidence.

Terminology is required for both disease-associated variant classes (e.g. missense variant), and the disease-associated consequence of those variants (e.g. altered gene product sequence). There may be good evidence for a particular variant class causing disease without a good understanding of the disease mechanism. For example, for *MYH7* and cardiomyopathy which is typically caused by missense variants, and less frequently splice variants, but not by PTCs: one might establish that disease requires the presence of an abnormal gene product, and the gene is not haploinsufficent, long before determining exactly how the beta myosin heavy chain protein function is perturbed (activating variants via loss of certain interactions; loss of ability to enter low energy relaxed state; gain of ATPase activity; etc.), or which particular perturbations lead to clinically distinct types of cardiomyopathy (hypertrophic cardiomyopathy vs dilated cardiomyopathy).

The 2015 ACMG/AMP sequence variant interpretation guideline provided a framework for classifying variants based on several evidence criteria indicative of benign or pathogenic features, including a criterion (PVS1) specific to predicted LoF variants. PVS1 is defined as ‘null variant (nonsense, frameshift, canonical ±1 or 2 splice sites, initiation codon, single or multi-exon deletion) in a gene where LoF is a known mechanism of disease.’ This is a somewhat oversimplified functional interpretation of a specific set of sequence variants. While these variant classes are indeed most likely to cause NMD and effectively be null or lead to LoF due to a truncated product, there is also potential for gain of function (GoF) through loss of a regulatory region (either terminal in the case of a truncation or internal in the case of an in-phase deletion), or action as a dominant negative or poison peptide. Furthermore a missense variant, not included in the list above, also has potential to act as LoF, GoF or as a dominant negative. The functional (and thus clinical) consequences of a given variant are only partially predictable from sequence alone. As such it is necessary to describe which predictable consequences have been associated with disease-gene pair, and/or are consistent with a known molecular mechanism of disease, if we are to apply appropriate filters in a variant-prioritization pipeline.

For a novel or previously uncharacterized variant, we usually have understanding of sequence consequence, e.g. amino acid sequence, but not (directly) the functional effect which speaks to mechanism. The ability to capture high level predictable consequences (disease-associated variant consequence) when the precise functional effect is unknown would be beneficial. For example a nonsense variant could lead to ‘decreased gene product level’, and a missense variant could lead to an ‘altered gene product sequence’ (altered amino acid sequence). Similarly, recognizing the high-level consequences of known pathogenic variants for a disease-gene pair reported in the literature or in ClinVar can help predict both mechanism and other variant classes that may have similar consequences. For example, if nonsense variants are pathogenic, we might expect other variants leading to ‘decreased gene product level’ such as frameshift variants to have similar effects, even if not previously observed. The weight given to PVS1 in the ACMG/AMP framework highlights the importance of correctly identifying disease associated variant consequences.

The terminology presented here is intended to be compatible with Ensembl Variant Effect Predictor (VEP) and other variant annotation tools that use Sequence Ontology (SO) terms for consequence. The SO is a structured, controlled vocabulary for the definition of sequence features used in biological sequence (e.g. DNA, RNA or peptide) annotation ^21^. There are ∼200 SO terms for variant consequence including a mix of terms to describe variant class (e.g. missense_variant SO:0001583) and variant function (e.g. gain_of_function_variant SO:0002053); with a subset of 33 variant classes used by Ensembl VEP, 42 by SnpEff ^26^, whereas Annovar ^27^ outputs 19 variant classes but does not use SO terms ^28^. Ensembl VEP requires outputs to be calculable based on sequence alone.

It was agreed to collaborate with SO to update the 33 variant consequence terms used by VEP. Redundant SO terms (e.g. downstream_gene_variant) were culled and new terms added to capture the different impact of variants either triggering or escaping nonsense-mediated-decay (NMD) (Table 2).

**Table 2.**
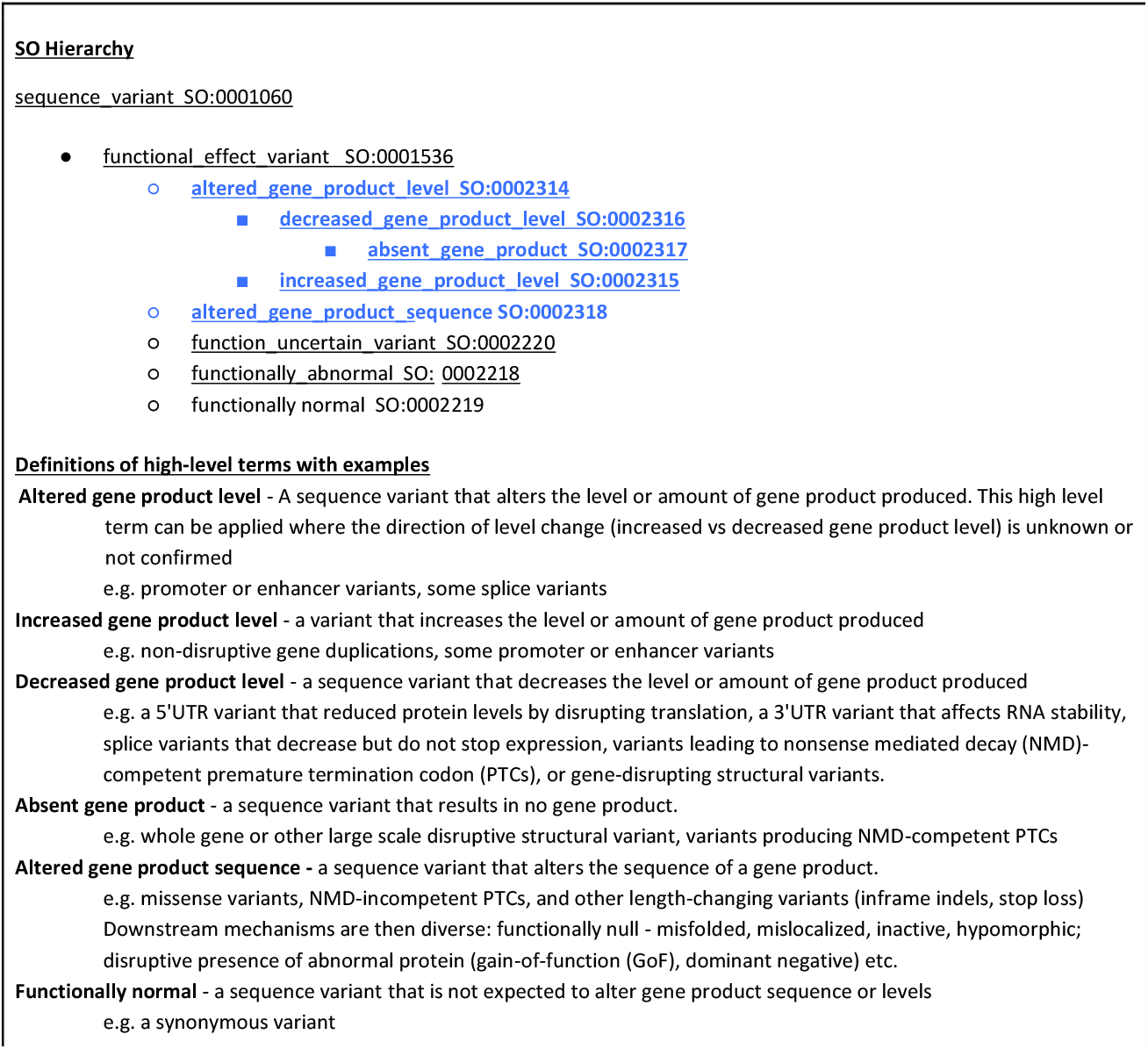
Hierarchy of SO disease-associated variant consequence terms. New high-level terms shown in blue.

High level terms to describe predictable variant consequences (disease-associated variant consequence terms) were proposed (table 2) and trialed by mapping consequence terms to SO variant class terms. Following iterative discussion and survey, a matrix was generated mapping six high-level predictable disease-associated variant consequence terms (altered gene product level, decreased gene product level, absent gene product, increased gene product level, altered gene product sequence, and functionally normal) to more specific variant classes described by SO variant class terms (Table 3). These are mapped via a semi-quantitative scale representing the likelihood of each consequence (1:almost never, 2:unlikely, 3:possible, 4:probable, 5:almost always), characterized from first principles by expert evaluation.

**Table 3.**
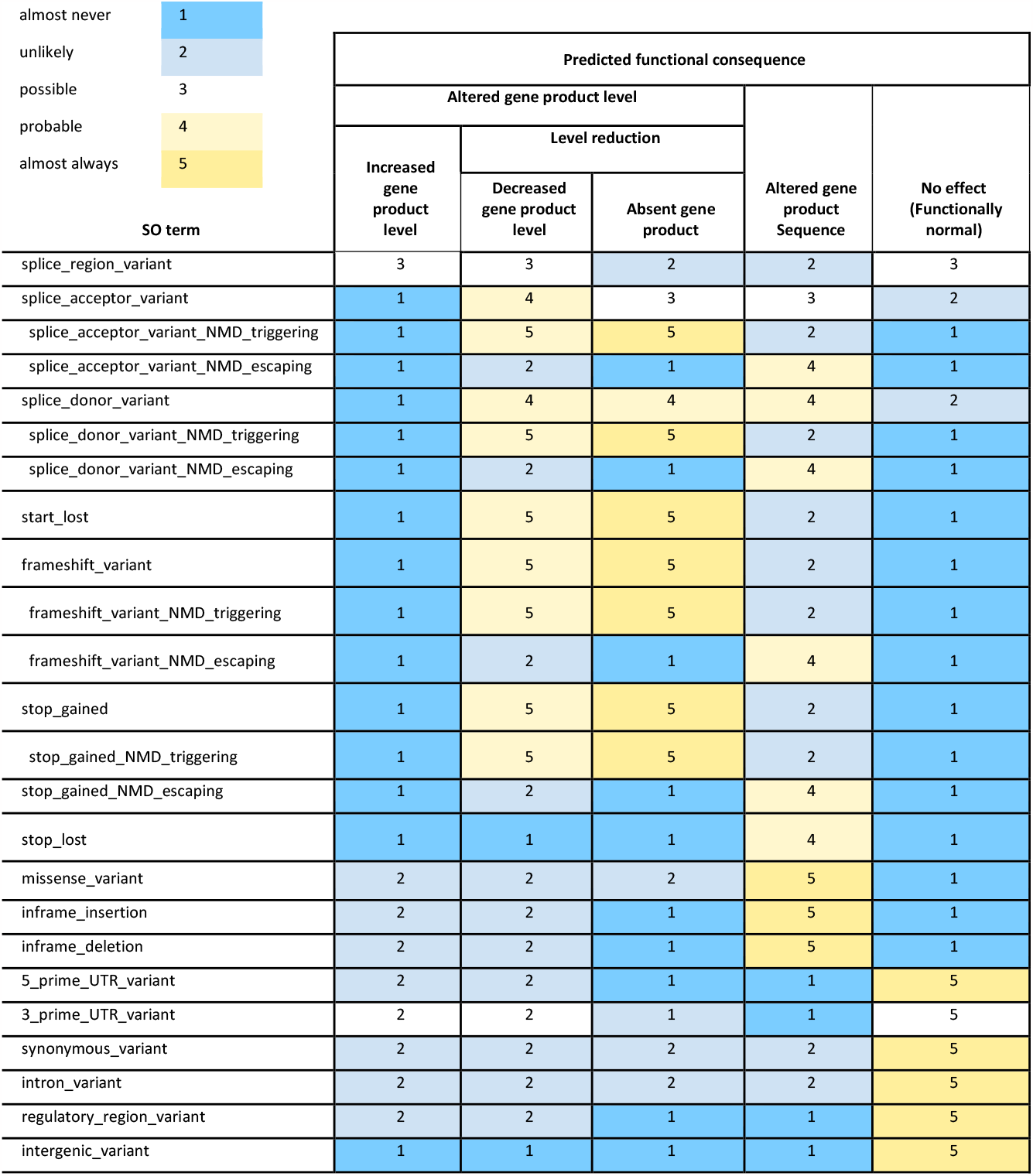
Matrix of six new high-level predicted functional consequences mapped to SO structural consequence terms via a semi-quantitative scale indicating likelihood of each high-level consequence. The semi-quantitative scale is characterized from first principles by expert evaluation.

In brief, different classes of variant may alter the level (abundance) or the sequence of the gene product, or may have no effect on either. A gene for our purpose is a segment of DNA that encodes an RNA that in turn performs some function; the product of a coding gene is a protein, and the product of a non-coding gene is non-coding RNA (ncRNA). For variants that alter gene product level, the direction of effect may be known (increased or decreased/absent), or unknown (altered). ‘Decreased gene product level’ therefore captures the group of variant classes typically analyzed together because they often result in a premature termination codon (PTC) - i.e. NMD-competent nonsense and frameshift variants, as well as most variants at canonical splice donor/acceptor sites - also sometimes described in the literature as (predicted) protein truncating variants (PTVs), or loss-of-function variants (LOF) (although missense variants can also result in decreased or absent protein levels through a variety of mechanisms). Metrics such as constraint have improved understanding of the impact of other, non-PTC, variants such as promoter or UTR variants on the level of gene product produced and these could also be captured by ‘Decreased gene product level’.

‘Altered gene product sequence’ captures sequence-altering variants such as missense, NMD-incompetent PTCs, and other length-changing variants (e.g. inframe indels, stop loss) that would change the amino acid sequence of a protein, or the nucleotide sequence of a ncRNA. Multiple disease-associated variant consequence terms can be linked to a given variant class, for example a splice donor variant may lead a ‘decreased gene product level’ and/or an ‘altered gene product sequence’ (altered amino acid sequence due to altered splicing).

When gathering evidence for a new disease-gene relationship it is likely that the types of variants associated with disease may be determined before their precise mechanistic consequences are understood. For example, a new gene-disease relationship may be curated with evidence that the disease-associated variant classes are those leading to ‘altered gene product sequence’, without knowing whether these lead to loss-of-function (e.g. through mis-folding, loss of an active site, or altered trafficking), or gain-of-function (increased enzyme activity or ion channel conductance, or new poison peptide activity). The more information available about a variant (e.g. computational, transcriptomic or functional studies), the more specific one can be with the functional consequence term, for example a 5’ UTR variant could lead to either increased or decreased expression and so in the absence of more specific knowledge the ‘altered gene product level’ term could be applied, but following expression studies this could be manually updated to either ‘increased gene product level’ or ‘decreased gene product level’.

### Results from Pilot study

We piloted the new terminology for inheritance, allelic requirement and disease mechanism using the ACMG SF v2.0, 59 genes (66 gene-disease pairs) ^22^. We generated curations for the purpose of piloting the terminology and these curations are not intended for application to clinical practice. The list includes genes related to inherited cardiovascular disease, cancer phenotypes and inborn errors of metabolism, and the gene disease pairs are characterized by a range of inheritance types and different disease mechanisms. All 66 gene disease pairs were successfully described using the new terminology (see supplemental data). Note that these are informal curations for the purpose of this study only; for official curations please see the ClinGen website https://clinicalgenome.org/.

Several changes were suggested and implemented following the pilot (See Supplementary file 1 for the initial draft terminology for comparison with final terminology in Tables 1 b&c). The inheritance modifier ‘Typified by age-related onset’ was added to allow description of conditions in which age of onset is typically later in life, i.e. adulthood, and in which penetrance is dependent on the age of the subject, this was then expanded to include the distinction between conditions which are typically congenital (e.g. congenital heart disease or cleft lip/palate), and those which present at any stage after birth (Table 1b). The term ‘Congenital onset’ was added. Child terms for age-related onset: ‘typically infantile onset’ ‘typically childhood onset’ and ‘typically adult onset’, were added to provide granularity where that information is known. Any combination of the age-related onset terms can be used: e.g. for a condition such as NF1 which typically presents either congenitally or in childhood, both the ‘Congenital Onset’ and ‘Typically childhood onset’ terms can be applied; alternatively for a condition such as dilated cardiomyopathy due to *TTN* which typically presents in adulthood, less frequently in adolescence on careful family screening, but not in infancy^29^, the terms ‘typically adult onset’ could be applied. This was found to be particularly relevant for the cancer susceptibility and inherited cardiovascular condition genes included in the pilot. The need to capture information regarding structural variants as well as sequence variants was also noted, and SO terms will be developed for these as a separate initiative.

Based on our experience of undertaking the pilot curations and incorporating the finalized terminology we have developed a suggested template for curation using this framework (see supplementary file 1).

## Discussion

With the increasing availability of DNA sequencing, application in clinical practice has become routine, including increasingly comprehensive sequencing with larger panels of genes, whole exome (WES), or whole genome (WGS) sequencing^30,31^. Consequently, the number of variants identified in patients undergoing genetic testing is increasing. Curation of gene-disease relationships and of inheritance pattern, allelic requirement, pathogenic variant classes, and disease-associated variant consequence are vital for the efficient and accurate classification of variants, and effective clinical application of genetic information ^6,16,32^.

Streamlined filtering systems that reduce the number of extraneous variants (i.e. benign or non-actionable) for a laboratory to manually review are of increasing importance with the convergence of routine genomic sequencing and guidelines for the reporting of secondary findings ^33^. The current responsibility for interpreting the clinical relevance and actionability of a variant falls to the reporting laboratory and ultimately the clinician. Some laboratories report all variants including VUS, while others only report pathogenic or likely pathogenic results. Secondary findings, by definition, are much more likely to fall outside the area of expertise of the reporting laboratory or clinician. For example, an oncologist or cancer geneticist may order WES or WGS from a specialist cancer genetics laboratory for their patient with suspected hereditary cancer but sequencing may identify a reportable secondary finding in a cardiovascular gene such as *KCNQ1* causing Long QT syndrome (LQTS) or *FBN1* causing Marfan Syndrome, and vice versa. Robust, structured data on disease mechanisms is key to accurate interpretation of potential significance.

We successfully engaged a diverse group of experts to establish consensus standard terms and a systematized approach for mode of inheritance, allelic requirement, and disease-associated variant consequence, and present these as a structured resource. The final terms presented in table 3 will be used by members of the GenCC in sharing gene curations, and when filtering variants for disease-relevant variant consequences. We suggest these terms may provide a standard terminology across diverse areas of clinical genetics, including clinical genetic laboratory reporting and gene-disease curation efforts.

This structured resource can be used by individual laboratories or curation programs alongside variant filtering pipelines. Following assessment of which variant classes are consistent with a disease using this framework, pipelines can then be adjusted to prioritize only those relevant classes.

Use of these standardized terms will facilitate assessment of which variant classes are likely to be disease-relevant (e.g. to apply PVS1), and evaluation of functional evidence, to determine which ACMG/AMP rules are applicable for a variant under interpretation. In addition to streamlining disease-gene curation and facilitating interpretation of variants in established disease-gene pairs, this standardized terminology will aid in assessment of novel potential disease-gene or disease-variant relationships. For a previously unseen variant, we usually understand the genetic consequence, but not (directly) the functional effect. Similarly we may not know the precise mechanism for a disease or variant, but can interpret likely disease-relevant variant classes.

One aim of the terminology is to aid interpretation of novel variants based on established pathogenic variant classes for a given gene-disease pair. In our pilot, curators were asked to test this utility by matching reported variant classes to high-level disease-associated variant consequence terms in the matrix (Table 3) and identifying additional variant classes that were likely to have the same disease-associated variant consequence. For example, if most known pathogenic variants for disease A were ‘stop gained’ (aka nonsense) which has a likelihood score of ‘5:almost always’ for ‘decreased gene product level’ and ‘absent gene product’, our pilot curators could then infer that a novel variant of a class that also has a high likelihood (4:probable or 5:almost always) for ‘decreased gene product level’ or ‘absent gene product’ (e.g. an NMD competent frameshift), could reasonably be included in filters for relevant variants. Additional variant classes identified in this way as potentially disease-causing will be dependent on the initial disease-causing variant classes identified. During our pilot it was noted that the output of curations of inheritance, allelic requirement, disease-associated variant consequence and predicted disease-causing variant class were curator-dependent. The variant classes recorded as pathogenic for a given disease-gene pair were sometimes dependent on how exhaustive a literature search was undertaken and the level of evidence each curator required in order to assign pathogenicity. Standardizing curation methods is beyond the scope of this project that instead focuses on providing a terminology framework for future work to be built upon.

For some gene-disease pairs there are well established, often recurrent, pathogenic variants that are the only pathogenic variant in that gene, or that are rare and atypical causes of disease. For example, almost all evidence for *KCNQ1* as a cause of short QT syndrome is derived from a single missense variant (p.V141M) via a gain of function mechanism^34^ but this does not inform whether other missense variants may cause disease or if LOF is also a possible mechanism of disease. While the proposed terminology provides a robust framework, additional structured data will be needed to fully represent the repertoire of disease-associated variation for any given gene.

For some disease-gene pairs such as malignant hyperthermia (*CACNA1S* and *RYR1*) the mechanism (e.g. dominant negative or haploinsufficiency) is not clear^35^. If there is only limited evidence for haploinsufficiency one should not assume that all variant classes predicted to reduce gene product would be pathogenic. The decision whether to retain only variants with a high likelihood of pathogenicity, or all variants that could plausibly have an effect consistent with pathogenesis of a particular disease, will depend on the desired sensitivity and specificity of a genomic interpretation pipeline.

Craniofrontonasal dysplasia due to *EFNB1*, an X-linked condition which does not manifest (fully) if hemizygous^23^ is an example of an edge case that required the introduction of a specific cross cutting inheritance modifier (requires heterozygosity HP:0034343). Every effort has been made to identify other edge cases through regular meetings of the consensus development panel and the terminology pilot, however it may become apparent that additional terms are necessary after this framework is more broadly applied.

Duchenne and Becker muscular dystrophy due to variants in dystrophin was also explored as an edge case as complex rearrangements in the gene are often encountered^25^. Approximately 60% of pathogenic dystrophin variants are large insertions or deletions that lead to frameshift errors downstream, whereas approximately 40% are substitutions or small insertions and deletions. Due to the structure and function of the protein even a large in-frame deletion may result in the milder Becker phenotype if the N and C termini of the protein remain intact. In addition, for many of the cancer predisposition genes, whole gene and other large deletions, and structural rearrangements can cause disease. Together these cases highlight the need for an ontology to describe structural variants which is currently under development as a separate GenCC initiative.

In summary, correctly classifying variants is of utmost importance for management of genetic conditions, including for family cascade testing and the delivery of screening and treatment services as well as supporting reproductive choices. Curation of gene-disease validity and of mode of inheritance, allelic requirement, and disease-associated variant consequences are vital to support accurate variant classification. Currently, groups, including academic and healthcare centers, private companies, and consortia, utilize different terminology to describe these three characteristics. The considerable discrepancy in the derivation and application of these terms generates confusion and risks discordant assertions about pathogenicity. The GenCC promotes use of standardized terms for structured representation of gene-disease relationships, including strength of evidence for the gene-disease association, disease mode of inheritance, allelic requirement, structural and functional consequences of genetic variation, and mechanism of pathogenicity. Here, we propose consensus terminology to aid in the characterization of gene-disease relationships. This will allow for harmonization across genetic resources, and aid variant curation, classification, and reporting.

## Data Availability

All data produced in the present work are contained in the manuscript, supplement and online at https://github.com/ImperialCardioGenetics/ACMGSF_pilot_curation/

## Acknowledgments

This work was supported by the Sir Jules Thorn Trust [21JTA], Wellcome Trust [107469/Z/15/Z; 200990/A/16/Z; 222883/Z/21/Z], Medical Research Council (UK), British Heart Foundation [RE/18/4/34215; FS/CRLF/21/23011], NHLI Foundation Royston Centre for Cardiomyopathy Research, and the NIHR Imperial College Biomedical Research Centre. MTD was supported by the National Human Genome Research Institute grant U24HG006834. JSB was supported by the National Human Genome Research Institute grant U24HG009650. FC was supported by the European Molecular Biology Laboratory. JF and DP were supported by Wellcome [grant number WT223718/Z/21/Z and 220540/Z/20/A Wellcome Sanger Institute Quinquennial Review 2021-2026]. CLM was supported by the National Human Genome Research Institute grant U24HG006834. ERR was supported by the National Human Genome Research Institute grant U24HG006834. PNR was supported by the National Human Genome Research Institute grant U24HG011449. David van Sant was funded by NIH: T15LM007124. PanelApp Australia is funded by Australian Genomics (NHMRC grants GNT1113531 and GNT2000001). NW is supported by a Sir Henry Dale Fellowship jointly funded by the Wellcome Trust and the Royal Society (220134/Z/20/Z) and the Rosetrees Trust (PGL19-2/10025). HLR was supported by the National Human Genome Research Institute grant U24HG006834. Open Targets is supported by Open Targets. The work performed by authors at European Molecular Biology Laboratory-European Bioinformatics Institute for the Transforming Genomic Medicine Initiative project was supported by the Wellcome Trust (WT200990/Z/16/Z).

For the purpose of open access, the authors have applied a CC BY public copyright license to any Author Accepted Manuscript version arising.

The views expressed in this work are those of the authors and not necessarily those of the funders.

## References

1. Chong, J.X. et al. The Genetic Basis of Mendelian Phenotypes: Discoveries, Challenges, and Opportunities. Am J Hum Genet 97, 199–215 (2015).

2. Richards, S. et al. Standards and guidelines for the interpretation of sequence variants: a joint consensus recommendation of the American College of Medical Genetics and Genomics and the Association for Molecular Pathology. 17, 405–24 (2015).

3. Riggs, E.R. et al. Technical standards for the interpretation and reporting of constitutional copy-number variants: a joint consensus recommendation of the American College of Medical Genetics and Genomics (ACMG) and the Clinical Genome Resource (ClinGen). Genet Med 22, 245–257 (2020).

4. Bean, L.J.H. et al. Diagnostic gene sequencing panels: from design to report-a technical standard of the American College of Medical Genetics and Genomics (ACMG). Genet Med 22, 453–461 (2020).

5. Strande, N.T. et al. Evaluating the Clinical Validity of Gene-Disease Associations: An Evidence-Based Framework Developed by the Clinical Genome Resource. Am J Hum Genet 100, 895–906 (2017).

6. Riggs, E.R. et al. Clinical validity assessment of genes frequently tested on intellectual disability/autism sequencing panels. Genet Med 24, 1899–1908 (2022).

7. Royer-Pokora, B. et al. Cloning the gene for an inherited human disorder--chronic granulomatous disease--on the basis of its chromosomal location. Nature 322, 32–8 (1986).

8. Claussnitzer, M. et al. A brief history of human disease genetics. Nature 577, 179–189 (2020).

9. Jabbari, J. et al. New exome data question the pathogenicity of genetic variants previously associated with catecholaminergic polymorphic ventricular tachycardia. Circ Cardiovasc Genet 6, 481–9 (2013).

10. Walsh, R. et al. Reassessment of Mendelian gene pathogenicity using 7,855 cardiomyopathy cases and 60,706 reference samples. Genet Med (2016).

11. Piton, A., Redin, C. & Mandel, J.L. XLID-causing mutations and associated genes challenged in light of data from large-scale human exome sequencing. Am J Hum Genet 93, 368–83 (2013).

12. Karczewski, K.J. et al. The ExAC browser: displaying reference data information from over 60 000 exomes. Nucleic Acids Res 45, D840–d845 (2017).

13. Karczewski, K.J. et al. The mutational constraint spectrum quantified from variation in 141,456 humans. Nature 581, 434–443 (2020).

14. DiStefano, M.T. et al. The Gene Curation Coalition: A global effort to harmonize gene-disease evidence resources. Genet Med 24, 1732–1742 (2022).

15. Ellingford, J.M. et al. Recommendations for clinical interpretation of variants found in non-coding regions of the genome. Genome Medicine 14, 73 (2022).

16. Rehm, H.L. et al. ClinGen--the Clinical Genome Resource. N Engl J Med 372, 2235–42 (2015).

17. Bamford, S. et al. The COSMIC (Catalogue of Somatic Mutations in Cancer) database and website. British Journal of Cancer 91, 355–358 (2004).

18. Fokkema, I.F. et al. LOVD v.2.0: the next generation in gene variant databases. Hum Mutat 32, 557–63 (2011).

19. Firth, H.V. et al. DECIPHER: Database of Chromosomal Imbalance and Phenotype in Humans Using Ensembl Resources. Am J Hum Genet 84, 524–33 (2009).

20. Köhler, S. et al. The Human Phenotype Ontology in 2021. Nucleic Acids Res 49, D1207–d1217 (2021).

21. Eilbeck, K. et al. The Sequence Ontology: a tool for the unification of genome annotations. Genome Biol 6, R44 (2005).

22. Kalia, S.S. et al. Recommendations for reporting of secondary findings in clinical exome and genome sequencing, 2016 update (ACMG SF v2.0): a policy statement of the American College of Medical Genetics and Genomics. Genet Med 19, 249–255 (2017).

23. Wieland, I. et al. Twenty-six novel EFNB1 mutations in familial and sporadic craniofrontonasal syndrome (CFNS). Hum Mutat 26, 113–8 (2005).

24. Amir, R.E. et al. Rett syndrome is caused by mutations in X-linked MECP2, encoding methyl-CpG-binding protein 2. Nat Genet 23, 185–8 (1999).

25. Muntoni, F., Torelli, S. & Ferlini, A. Dystrophin and mutations: one gene, several proteins, multiple phenotypes. The Lancet Neurology 2, 731–740 (2003).

26. Cingolani, P. et al. A program for annotating and predicting the effects of single nucleotide polymorphisms, SnpEff: SNPs in the genome of Drosophila melanogaster strain w1118; iso-2; iso-3. Fly (Austin) 6, 80–92 (2012).

27. Wang, K., Li, M. & Hakonarson, H. ANNOVAR: functional annotation of genetic variants from high-throughput sequencing data. Nucleic Acids Res 38, e164 (2010).

28. McLaren, W. et al. The Ensembl Variant Effect Predictor. Genome Biology 17, 122 (2016).

29. Fatkin, D. et al. Titin truncating mutations: A rare cause of dilated cardiomyopathy in the young. Progress in Pediatric Cardiology 40, 41–45 (2016).

30. Brittain, H.K., Scott, R. & Thomas, E. The rise of the genome and personalised medicine. Clin Med (Lond) 17, 545–551 (2017).

31. Manickam, K. et al. Exome and genome sequencing for pediatric patients with congenital anomalies or intellectual disability: an evidence-based clinical guideline of the American College of Medical Genetics and Genomics (ACMG). Genet Med 23, 2029–2037 (2021).

32. Thomson, K.L. et al. Analysis of 51 proposed hypertrophic cardiomyopathy genes from genome sequencing data in sarcomere negative cases has negligible diagnostic yield. Genet Med 21, 1576–1584 (2019).

33. Miller, D.T. et al. ACMG SF v3.1 list for reporting of secondary findings in clinical exome and genome sequencing: A policy statement of the American College of Medical Genetics and Genomics (ACMG). Genet Med 24, 1407–1414 (2022).

34. Lee, H.C. et al. Pro-arrhythmogenic Effects of the V141M KCNQ1 Mutation in Short QT Syndrome and Its Potential Therapeutic Targets: Insights from Modeling. J Med Biol Eng 37, 780–789 (2017).

35. Johnston, J.J. et al. Updated variant curation expert panel criteria and pathogenicity classifications for 251 variants for RYR1-related malignant hyperthermia susceptibility. Human Molecular Genetics 31, 4087–4093 (2022).

