## Supplementary table S1 Supplementary table S2 Supplementary table S3 Supplementary information 1 Supplementary information 2 for "Towards robust clinical genome interpretation: developing a consistent terminology to characterize disease-gene relationships - allelic requirement, inheritance modes and disease mechanisms"

**Supplementary table S1**: **Table showing alignment of inheritance terms used by GenCC founder members at initial meeting**

Abbreviations:

GDM - Gene Disease Map (https://doi.org/10.17605/OSF.IO/S4PVA)

GHR - Genetics Home Reference (merged into MedlinePlus as of October 1, 2020)

DDG2P - The Development Disorder Genotype - Phenotype Database

EMBL-EBI **-** European Molecular Biology Laboratory European Bioinformatics Institute

HPO – Human Phenotype Ontology

| **Harmonization** | **ClinGen** | **GDM** | **GHR** | **Orphanet** | **DDG2P-Decipher** | **EMBL-EBI HPO term** |
|---|---|---|---|---|---|---|
| Autosomal dominant | Autosomal Dominant | Autosomal Dominant | Autosomal Dominant | Autosomal Dominant | Monoallelic | Autosomal dominant inheritance |
| Optional terms? | *optional specification:* with maternal/paternal imprinting, sex-limited, with genetic anticipation, primarily or exclusively de novo | | | | Imprinted | Autosomal dominant contiguous gene syndrome<br>Autosomal dominant germline de novo mutation<br>Autosomal dominant inheritance with maternal imprinting<br>Autosomal dominant inheritance with paternal imprinting<br>Autosomal dominant somatic cell mutation<br>Sex-limited autosomal dominant<br>Male-limited autosomal dominant<br>Semidominant mode of inheritance |
| Autosomal Recessive | Autosomal Recessive | Autosomal Recessive | Autosomal Recessive | Autosomal Recessive | Biallelic | Autosomal recessive inheritance |
| Optional terms? | *optional specification:* sex-limited, with genetic anticipation | | | | | Sex-limited autosomal recessive inheritance |
| | | | | | | Gonosomal inheritance |
| X-linked | X-linked | X-linked Dominant | X-linked | | | X-linked inheritance |
| *optional specification:* | *optional specification:* | | OR | | | |
| dominant, recessive, | dominant, recessive, | | X-Linked Dominant | X-linked dominant | X-linked dominant | X-linked dominant inheritance |
| primarily recessive with milder female expression | primarily recessive with milder female expression | | OR | | | ? X-linked semi dominant |
| | | X-linked Recessive | X-linked Recessive | X-linked recessive | Hemizygous | X-linked recessive inheritance |
| Y-linked | Y-linked needs to be added | Y-linked | Y-linked | Y-linked | | Y-linked inheritance |
| Mitochondrial | Mitochondrial | Mitochondrial | Mitochondrial | Mitochondrial | Mitochondrial | Mitochondrial inheritance |
| Complex | | Complex | | Multigenic/ multifactorial | Digenic | Multifactorial inheritance<br>Digenic inheritance<br>Oligogenic inheritance<br>Polygenic inheritance |
| | | | | | Mosaic | Somatic mutation<br>Somatic mosaicism |

**Supplementary table S2**: **Table showing alignment of allelic requirement terms used by GenCC founder members at initial meeting.**

Note not all founder members had separate allelic requirement terms in addition to inheritance terms.

Abbreviations:

PAR - pseudoautosomal region.

G2P - Gene2Phenotype

| **Proposal** | **Decipher (single variant annotation)** | **G2P** | **Genomics England PanelApp** |
|---|---|---|---|
| monoallelic_autosomal | Heterozygous | Monoallelic | Monoallelic, autosomal<br>(maternal/paternal imprinting) |
| biallelic_autosomal | Homozygous (Compound het does not exist) | Biallelic | Biallelic, autosomal |
| | | | Both<br>*Either: BOTH monoallelic and biallelic, autosomal or pseudoautosomal*<br>*Or*<br>*BOTH monoallelic and biallelic, autosomal or pseudoautosomal (but BIALLELIC mutations cause a more SEVERE disease form), autosomal or pseudoautosomal* |
| monoallelic_X_heterozygous | | X-linked Dominant | X linked: hemizygous in males, monoallelic in females<br>*X linked: hemizygous mutation in males, monoallelic mutations in females may cause disease (may be less severe, later onset than males)* |
| monoallelic_X_hemizygous | Hemizygous | Hemizygous | X linked: hemizygous in males, Biallelic in females |
| Mitochondrial | | Mitochondrial | Mitochondrial |
| | | Digenic | We use a tag for this, rather than it being collected in the mode of inheritance field |
| monoallelic_Y_hemizygous | | | |
| monoallelic_PAR | | | |
| biallelic_PAR | | | |

**Supplementary table S3**: **Table showing redundant HPO inheritance terms at initial review**
Abbreviations: HPO - Human phenotype ontology

| |
|---|
| Redundant HPO inheritance terms<br>• Sporadic<br>• Heterogeneous |
| HPO inheritance terms converted to Inheritance modifier terms<br>• Uniparental disomy<br>• Genetic anticipation<br>• Contiguous gene syndrome |
| Child terms depreciated after creation of Inheritance Modifier HPO category<br>• Sex-limited autosomal dominant<br> ○ Male-limited autosomal dominant<br>• Autosomal dominant germline de novo mutation<br>• Autosomal dominant inheritance with paternal imprinting<br>• Autosomal dominant somatic cell mutation<br>• Autosomal dominant inheritance with maternal imprinting<br>• Autosomal dominant contiguous gene syndrome<br>• Sex-limited autosomal recessive inheritance<br>• Autosomal dominant contiguous gene syndrome |

**Supplementary information**

**Supplementary information 1. Early iteration of terminology used for pilot curation of gene-disease relationships for ACMG SF 2.0 genes**

NB This document includes initial terminology given to pilot curators for the use of standardised terms for gene-disease pairs. The terminology was iteratively modified and improved based on feedback from pilot curators.

**Inheritance:**
High level terms and (optional) modifiers:

**High level terms**

- Autosomal Dominant
  - with maternal/paternal imprinting
  - with sex-limited expression
  - with genetic anticipation
- Autosomal Recessive
  - with sex-limited expression
  - with genetic anticipation
- X-linked
  - Dominant
  - Strictly recessive
  - Primarily recessive (with milder female expression)
- Y-linked
- Mitochondrial
- PAR recessive
- PAR dominant

**Crosscutting modifiers:**

- De novo
- Mosaic
- Digenic
- Incomplete penetrance

Notes:

- Use of **cross cutting modifiers** enables recording of data important to reproductive advice and family screening.
  - **Mosaic:** in a situation where, for example, a constitutive mutation is lethal and a condition is exclusively or predominantly mosaic. Should not be used to describe the situation of mosaicism in a parent.
  - **De Novo**: description of conditions that are exclusively or predominantly de novo due to the limited reproductive fitness of affected individuals. Should not be used to describe a situation where a gene just has a high new mutation rate.

- **Digenic**: for example, in Long QT syndrome, heterozygosity for pathogenic variants in two different genes occurs more frequently than would be expected by chance. It is generally associated with a more severe phenotype and the possibility that both parents have pathogenic variants should be considered. The genes involved should be specified.
- **Incomplete penetrance**

**Allelic requirement:**
High level terms and (optional) modifiers:

| |
|---|
| **High level terms:**<br>monoallelic_autosomal<br>monoallelic_X_heterozygous<br>monoallelic_X_hemizygous<br>monoallelic_Y_ hemizygous<br>monoallelic_PAR<br>biallelic_PAR<br>biallelic_ autosomal |
| **Modifiers:**<br>Mosaic<br>Imprinted<br>Requires heterozygosity<br>Digenic |

Notes:

- An **X-linked dominant** condition would be curated as monoallelic_X_het and we would understand that those diseases manifest when het or hem (or indeed hom/compound het - though this may be more severe or lethal).
- An **X-linked recessive** condition would be curated as monoallelic_X_hem and would not manifest when heterozygous (though they can manifest with ameliorated phenotype, or manifest if skewed inactivation etc - we intend that this is implicit in the term, as characteristic of many sex-linked disorders, and do not anticipate that an additional modifier term is needed to communicate this, unless the het phenotype is sufficiently distinct as to be classified as a different disease entity)
- Terms are specific to each disease-gene pair, so for example if there is good evidence for manifesting carriers of X_Hem disorders presenting in infancy/early childhood that would be coded as X_Hem in DDG2P. If carriers only have late onset cardiomyopathy (e.g. female carrier of DMD) they would be X_het in the Cardiac panel but not DD.
- **Mosaic** is intended for conditions that are typically lethal when constitutive
- **Imprinted** requires that the abnormal allele be paternal or maternal in origin.
- **Requires heterozygosity** covers edge cases such as Craniofrontonasal dysplasia due to EFNB1 - which requires heterozygosity and would not manifest (fully) if hemizygous. Importantly the mutant allele can be inherited from a normal or very mildly affected father.

**Disease associated variant consequences:**
High level terms to describe variant consequences:

- Dose Change
  - Dose reduction
    - Decreased gene product level
    - Absent gene product
  - Increased gene product level
- Altered gene product sequence

Notes:

- **Dose reduction** – for example PTCs (protein truncating), gene-disrupting SVs, and gene-deletions (assuming NMD-competent PTC, and with caveats about splicing)
- **Increased gene product** – for example non-disruptive gene duplications, some promoter or enhancer variants
- **Altered gene product sequence** – for example NMD-incompetent PTCs, other length-changing variants (inframe indels, stop loss), and missense. Downstream mechanisms can be diverse: functionally null - misfolded, mislocalized, inactive, hypomorphic; disruptive presence of abnormal protein (gain of function, dominant negative etc)

### Variant class/ consequence matrix

| | |
|---|---|
| almost never | 1 |
| possible but unlikely | 2 |
| 50:50 | 3 |
| probable | 4 |
| almost always | 5 |

| | Predicted functional consequence | | | | | |
|---|---|---|---|---|---|---|
| | Altered gene product level | | | | | |
| | | | level reduction | | | |
| SO term | Unspecified gene product level | Increased gene product level | Decreased gene product level | *Absent gene product* | Altered gene product Sequence | No effect |
| splice_region_variant | 3 | 3 | 3 | 2 | 2 | 3 |
| splice_acceptor_variant | 2 | 1 | 4 | 3 | 3 | 2 |
| splice_donor_variant | 2 | 1 | 4 | 4 | 4 | 2 |
| start_lost | 1 | 1 | 4 | 5 | 2 | 1 |
| frameshift_variant | 1 | 1 | 4 | 5 | 2 | 1 |
| stop_gained | 1 | 1 | 4 | 5 | 2 | 1 |
| (stop_gained predicted to undergo NMD) | 1 | 1 | 4 | 5 | 2 | 1 |

| | | | | | | |
|---|---|---|---|---|---|---|
| (stop_gained predicted to escape NMD) | 1 | 1 | 2 | 1 | 4 | 1 |
| stop_lost | 1 | 1 | 1 | 1 | 4 | 1 |
| missense_variant | 2 | 2 | 2 | 2 | 5 | 1 |
| inframe_insertion | 2 | 2 | 2 | 1 | 5 | 1 |
| inframe_deletion | 2 | 2 | 2 | 1 | 5 | 1 |
| 5_prime_UTR_variant | 2 | 2 | 2 | 1 | 1 | 5 |
| (gain of upstream Start [uORF]) | 1 | 1 | 4 | 3 | 1 | 3 |
| (gain of upstream Start [oORF]) | 1 | 1 | 5 | 4 | 1 | 2 |
| 3_prime_UTR_variant | 3 | 3 | 3 | 2 | 1 | 3 |
| synonymous_variant | 2 | 2 | 2 | 2 | 2 | 5 |
| intron_variant | 2 | 2 | 2 | 2 | 2 | 5 |
| regulatory_region_variant | 2 | 2 | 2 | 1 | 1 | 5 |
| intergenic_variant | 1 | 1 | 1 | 1 | 1 | 5 |

**Supplementary information 2. Using a framework of standardised terminologies to define inheritance, allelic requirement, disease-associated variant classes, and disease-associated variant consequence for gene-disease pairs.**

This document provides a template and guidance for the curation of inheritance, allelic requirement and disease-associated variant consequences for gene-disease pairs already curated by ClinGen using standardised terminology.

**TEMPLATE**

**Review of evidence:**

**ClinGen**:
Include summary of evidence to support gene-disease relationship. This can be found at https://clinicalgenome.org/ and searching for the specific gene.
Paste web link for ClinGen evidence summary page here e.g. for KCNQ1 http://search.clinicalgenome.org/kb/genes/HGNC:6294.
If the summary page is not on the ClinGen website, these can sometimes be found in the Supplementary data of the ClinGen curation paper.

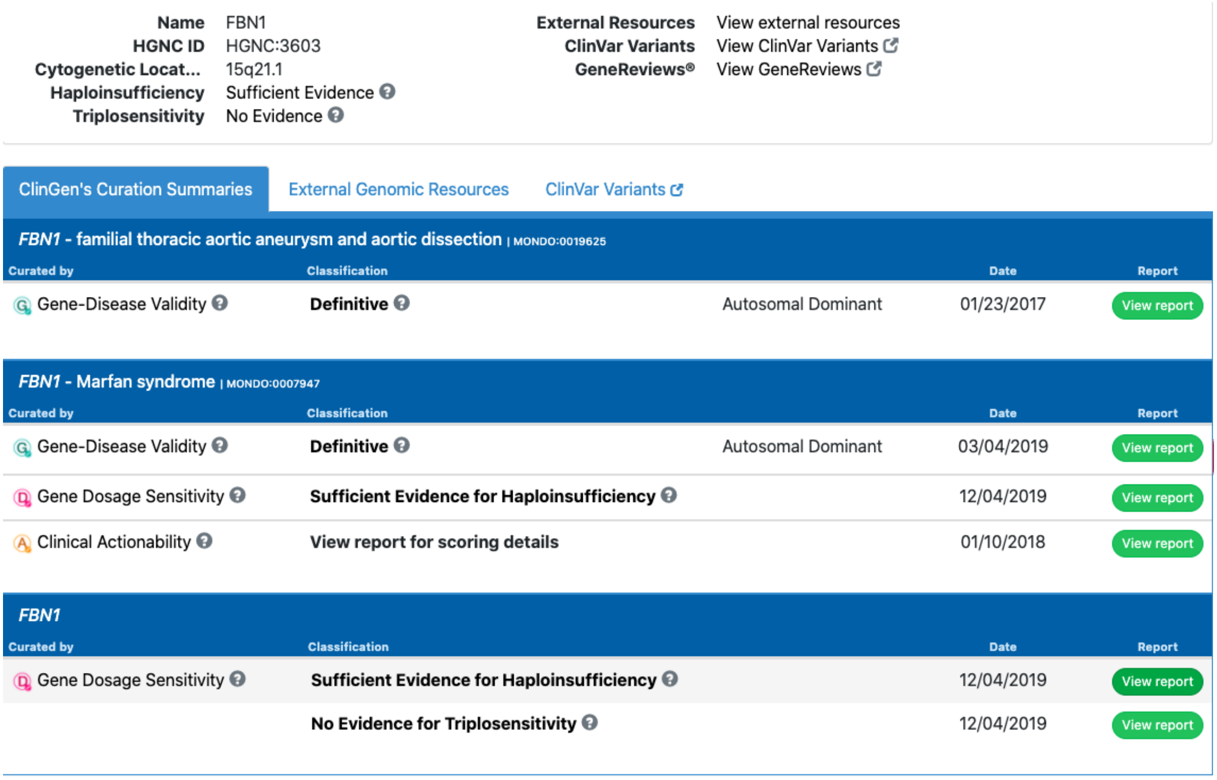

| | | | |
|---|---|---|---|
| **Name** | FBN1 | **External Resources** | View external resources |
| **HGNC ID** | HGNC:3603 | **ClinVar Variants** | View ClinVar Variants |
| **Cytogenetic Locat...** | 15q21.1 | **GeneReviews®** | View GeneReviews |
| **Haploinsufficiency** | Sufficient Evidence | | |
| **Triplosensitivity** | No Evidence | | |

ClinGen's Curation Summaries | External Genomic Resources | ClinVar Variants

*FBN1* - familial thoracic aortic aneurysm and aortic dissection | MONDO:0019625

| Curated by | Classification | | Date | Report |
|---|---|---|---|---|
| Gene-Disease Validity | **Definitive** | Autosomal Dominant | 01/23/2017 | View report |

*FBN1* - Marfan syndrome | MONDO:0007947

| Curated by | Classification | | Date | Report |
|---|---|---|---|---|
| Gene-Disease Validity | **Definitive** | Autosomal Dominant | 03/04/2019 | View report |
| Gene Dosage Sensitivity | **Sufficient Evidence for Haploinsufficiency** | | 12/04/2019 | View report |
| Clinical Actionability | **View report for scoring details** | | 01/10/2018 | View report |

*FBN1*

| Curated by | Classification | Date | Report |
|---|---|---|---|
| Gene Dosage Sensitivity | **Sufficient Evidence for Haploinsufficiency** | 12/04/2019 | View report |
| | **No Evidence for Triplosensitivity** | 12/04/2019 | View report |

**Review of source material:**
Review and include the reference (PMID) for the relevant ClinGen gene-disease validity paper e.g. for hypertrophic cardiomyopathy PMID: 30681346. This is likely to include useful summary information and publications to refer to.

**Other Literature review:**

This is to gather new information, not to re-evaluate the gene-disease relationship. Evidence is collected primarily from published peer-reviewed literature, but can also be present in publicly accessible resources, such as variant databases. Up-to-date reviews from centres with particular expertise in a given gene or disease are particularly helpful.

Useful publication search engines include:
PubMed
Google Scholar
LitVar
GeneCards
Mastermind

Other useful information
GeneReviews and the "Molecular Genetics" section
OMIM
ClinVar to search for relevant variant classes
PanelApp (If using a resource like PanelApp need to reference the assertion and check original references)

As these gene-disease pairs have all been classified as "Definitive" or "Strong" by ClinGen, they are well established and there may be abundant information. The goal is not to re-evaluate the gene-disease validity, and the literature review therefore does not have to be exhaustive. The literature search should be focused on establishing inheritance pattern, allelic requirement and where possible disease-associated variant class and functional consequences.

For example, for some gene-disease pairs it may be well established that the pattern of inheritance is autosomal dominant but there may be a small number of reports of recessive inheritance. A broad search of the literature can determine if other modes of inheritance have been reported, using search terms:
Gene AND disease AND ("recessive" OR "autosomal recessive" OR "homozyg*" OR "compound heterozyg*" OR "biallelic")
Gene AND disease AND (dominant OR "autosomal dominant" OR monoallelic OR heterozyg*)
Gene AND disease AND ("x-linked" OR "x linked" OR "X chromosome" OR "X linked dominant" OR "X linked recessive")

Any reports of a different mode of inheritance should be reviewed to see if they are relevant or not. For many genes, a second hit may lead to a more severe phenotype but that does not necessarily mean the inheritance follows a recessive or digenic pattern as both the first and second hit would in fact cause disease in isolation.

For disease-associated variant consequence and mechanism, literature review should focus on establishing the most likely consequences. Curators should review the evidence for haploinsufficiency in the ClinGen Dosage Sensitivity curation (http://search.clinicalgenome.org/kb/gene-dosage?page=1&size=25&order=asc&sort=symbol&search=), pathogenic/likely pathogenic variant classes on ClinVar (Simple ClinVar can be a helpful tool to search ClinVar, see screen shot below and link http://simple-clinvar.broadinstitute.org) and other public variant databases where available. For well described genes, recent publications re-evaluating variants, expert reviews, meta-analyses, and reviews of burden testing are highly relevant.

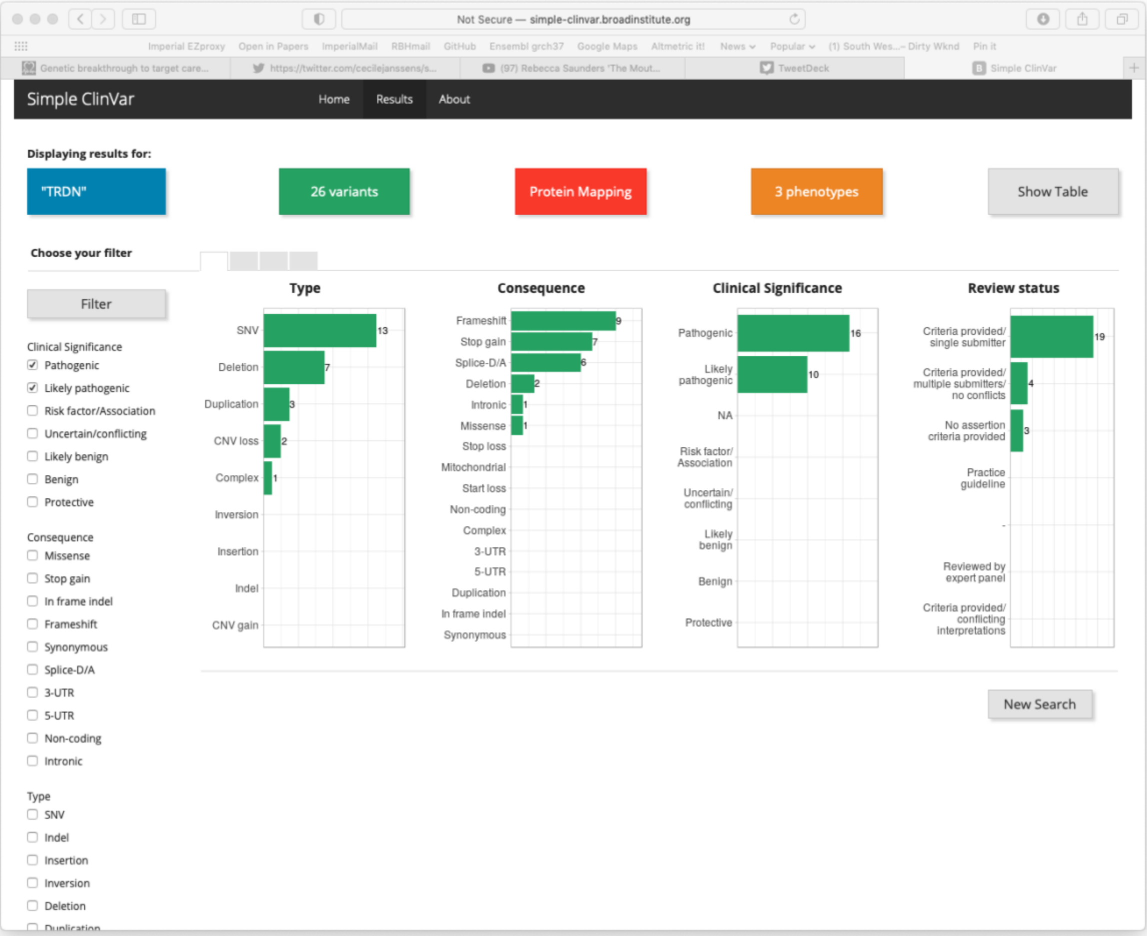

It is not necessary to review every variant. However, if for 12example the predominant class of variant is missense but there are a small number of nonsense mutations reported, extra time should be spent determining whether there is sufficient evidence to include these as a pathogenic variant class before expanding the disease mechanism. Sufficient evidence could include segregation or functional evidence.

If high level reviews are not available for a gene-disease pair, then a broad literature search may be necessary e.g. Gene AND disease AND (variant OR mutation). For a variant class to be included that would add to the predicted functional consequence, there should be sufficient qualitative evidence to support that such as segregation, functional or burden data.

Where there is uncertainty that cannot be resolved a note should be made in the narrative summary. Include PMIDs where possible and or links to other resources.

### Inheritance and Allelic Requirement

List inheritance and allelic requirement terms and any number of appropriate modifier terms.
Use of modifiers enables recording of data important to reproductive advice and family screening.

**Harmonised allelic requirement and Mendelian inheritance terms.**

Abbreviations: HPO - Human phenotype ontology, PAR - pseudoautosomal region.

| **allelic requirement term** | **inheritance term** | **HPO id** |
|---|---|---|
| monoallelic_autosomal | Autosomal Dominant | HP:0000006 |
| biallelic_autosomal | Autosomal Recessive | HP:0000007 |
| monoallelic_X_heterozygous | X-linked Dominant | HP:0001423 |
| monoallelic_X_hemizygous | X-linked Recessive | HP:0001419 |
| monoallelic_Y_hemizygous | Y-linked | HP:0001450 |
| mitochondrial | Mitochondrial | HP:0001427 |
| monoallelic_PAR | PAR dominant | HP:0034340 |
| biallelic_PAR | PAR recessive | HP:0034341 |

**Inheritance modifier terms**- these optional terms can be combined with either inheritance terms or allelic requirement terms to provide additional information about the relationship of a disease-gene pair.

| **Inheritance Modifier<br>HP:0034335<br>(Parent and child terms)** | **Definition<br>(Parent term)** |
|---|---|
| Typically mosaic | Description of conditions in which, for example, constitutive mutation is lethal, and cases are exclusively or predominantly mosaic. A much lower variant allele fraction (VAF) cut-off would be needed in analysis pipelines. |
| Typically *de novo*<br>HP:0025352 | Description of conditions that are exclusively or predominantly observed due to *de novo* variants. In some cases, this may be due to the limited reproductive fitness of affected individuals. |
| Typified by incomplete penetrance<br>HP:0003829<br>- Typified by moderate penetrance HP:4000159 | Description of conditions in which not all individuals with a given genotype exhibit the disease. Penetrance is the proportion that develop disease given a lifespan of 80 years.<br>Examples include, CYP1B1 glaucoma which has approximately 90% penetrance; Van der Woude syndrome due to *IRF6* causes cleft lip and/or palate with |

| | |
|---|---|
| - Typified by high penetrance HP:4000158 | penetrance estimated at 80%; *C9orf72* causes frontotemporal dementia and/or amyotrophic lateral sclerosis with approximately 50% penetrance. |
| Typified by complete penetrance HP:0003829 | Description of conditions in which all individuals with a given genotype exhibit the disease within a lifespan of 80 years. For example, penetrance of Neurofibromatosis type 1 due to *NF1* is close to 100%. |
| Variable age of onset | Description of conditions in which age of onset is highly variable and in which manifestation of the disease phenotype is not dependent on the age of the subject. |
| Typified by age-related onset HP:0003831<br>- Typically infantile onset<br>- Typically childhood onset<br>- Typically adult onset | Description of conditions in which age of onset is typically not congenital and in which manifestation of the disease phenotype is dependent on the age of the subject. |
| Congenital onset | Description of conditions which are manifest at or before birth, for example cleft lip or talipes. |
| Imprinted HP:0034338<br>- With maternal imprinting HP:0012275<br>- With paternal imprinting<br>- HP:0012274 | Requires that the abnormal allele be paternal or maternal in origin, depending on the disease-gene relationship. Imprinting refers to a normal developmental process in which either the paternal or maternal allele is inactivated, depending on the specific locus, thus leading to expression from only one copy of the gene. Disease typically manifests when a deleterious variant is inherited from a parent whose copy of the gene would normally be expressed, but not when a deleterious variant is inherited from a parent whose copy of the gene would normally be inactivated. |
| Displays anticipation HP:0003743 | A phenomenon in which the severity of a disorder increases, or the age of onset decreases, as the disorder is passed from one generation to the next, typically due to expansion of a repeat sequence. For example, Myotonic Dystrophy is caused by triplet repeat expansion in the *DMPK* gene. |
| Requires heterozygosity HP:0034343 | Covers rare instances of a condition that is most severe in the heterozygous state. Such disorders are rare and currently all are X-linked. Most X-linked recessive conditions manifest if hemizygous in males, or biallelic in females, though may have a mild phenotype in the heterozygous state in females. However, Craniofrontonasal dysplasia due to *EFNB1,* and *PCDH19*-related epilepsy, are both X-linked dominant and paradoxically more severe in females. Hemizygous males may be mildly affected but seldom manifest the full phenotype. Importantly the mutant allele can be inherited from a normal or very mildly affected father. The mechanism is currently accepted to be due to cellular interference whereby the two distinct cell populations (those with and without the variant) exhibit abnormal cellular interactions in the mosaic state - in women, who are functionally mosaic due to random X inactivation, or mosaic males. The same mechanism could theoretically be observed in autosomal genes with a mosaic variant. |
| Sex-limited expression HP:0001470 | Condition in which the phenotype only manifests in one sex, i.e. either manifests in males or females but not both. Example: Autosomal recessive sex reversal due |

| | |
|---|---|
| - Male-limited expression HP:0001475<br>- Female- limited expression<br>- HP:0034344 | to DHH on chr12 manifests only in XY males causing gonadal dysgenesis, while XX females are phenotypically normal. |
| Contiguous gene syndrome HP:0001466 | Syndrome caused by the effects of abnormality (typically a deletion or duplication) of 2 or more adjacent genes. |

**Notes:**

- **Mitochondrial -** the inheritance of a trait encoded in the mitochondrial genome. Persons with mitochondrial disease may be male or female but the mode of inheritance is strictly maternal. No male with the disease can transmit it to their offspring.
- **PAR** - genes within the pseudoautosomal regions (PAR) are inherited like autosomal genes. PAR1 comprises 2.6mb of the short-arm of both X and Y chromosomes in humans. PAR2 is at the tip of the long arms, spanning 320kb.
  Normal male mammals have two copies of these genes: one in the pseudoautosomal region of their Y chromosome, the other in the corresponding portion of their X chromosome. Normal females also possess two copies of pseudoautosomal genes, as each of their two X chromosomes contains a pseudoautosomal region. Crossing over between the X and Y chromosomes is normally restricted to the pseudoautosomal regions; thus, pseudoautosomal genes exhibit an autosomal, rather than sex-linked, pattern of inheritance. So, females can inherit an allele originally present on the Y chromosome of their father.
- For monoallelic_X_het (**X-linked dominant)** conditions, we would understand that those diseases manifest when het or hem (or indeed hom/compound het - though this may be more severe or lethal).
- For monoallelic_X_hem (**X-linked recessive)** conditions, we would understand that these would not manifest when heterozygous (though they can manifest with ameliorated phenotype, or manifest if skewed inactivation etc - primarily recessive with milder female expression)
- Terms are specific to each disease gene pair. Considering a hypothetical example of a gene on the X-chromosome in which biallelic or hemizygous monoallelic variation causes congenital structural heart abnormalities, but a heterozygous monoallelic variant typically presents with late onset cardiomyopathy, this might be coded as monoallelic_X_hemizygous for congenital heart disease, and appropriate filtering applied in a developmental disorders panel for diagnosis of an infant, and monoallelic_X_heterozygous (age-related onset) for cardiomyopathy, with different variant filtering applied for a cardiac gene panel analysis in an adult. This has the advantage of tracing the evidence for each disease association.

**<u>Disease-associated variant classes:</u>**

**List variant classes (**SO terms**) in this gene proven to cause this disease:**
Consider whether the disease is associated with:
- missense & in frame variants
- Protein terminating codon (PTCs) (premature truncating variants (PTV)) or loss of function (LoF) or radical variants

for PTCs need to consider whether nonsense mediated decay (NMD) competent or not

In practice it is useful to know whether a gene-disease pair is associated with missense only, truncating only, or both)
See matrix below for variant classes and SO terms

**List other variant classes predicted to lead to the same functional consequence:**
Other variant classes that could be predicted to lead to the same functional consequence based on inferred mechanism (score 4 or 5, see matrix below) and therefore might cause the same phenotype.

Matrix of six new high-level predicted functional consequences mapped to SO structural consequence terms via a semi-quantitative scale indicating likelihood of each high-level consequence

The semi-quantitative scale is characterized from first principles by expert evaluation.

| | |
|---|---|
| almost never | 1 |
| unlikely | 2 |
| possible | 3 |
| probable | 4 |
| almost always | 5 |

| SO term | Predicted functional consequence | | | | |
|---|---|---|---|---|---|
| | Altered gene product level | | | Altered gene product Sequence | No effect (Functionally normal) |
| | Increased gene product level | Level reduction | | | |
| | | Decreased gene product level | Absent gene product | | |
| splice_region_variant | 3 | 3 | 2 | 2 | 3 |
| splice_acceptor_variant | 1 | 4 | 3 | 3 | 2 |
| splice_acceptor_variant_NMD_triggering | 1 | 5 | 5 | 2 | 1 |
| splice_acceptor_variant_NMD_escaping | 1 | 2 | 1 | 4 | 1 |
| splice_donor_variant | 1 | 4 | 4 | 4 | 2 |
| splice_donor_variant_NMD_triggering | 1 | 5 | 5 | 2 | 1 |
| splice_donor_variant_NMD_escaping | 1 | 2 | 1 | 4 | 1 |
| start_lost | 1 | 5 | 5 | 2 | 1 |
| frameshift_variant | 1 | 5 | 5 | 2 | 1 |
| frameshift_variant_NMD_triggering | 1 | 5 | 5 | 2 | 1 |
| frameshift_variant_NMD_escaping | 1 | 2 | 1 | 4 | 1 |
| stop_gained | 1 | 5 | 5 | 2 | 1 |
| stop_gained_NMD_triggering | 1 | 5 | 5 | 2 | 1 |
| stop_gained_NMD_escaping | 1 | 2 | 1 | 4 | 1 |
| stop_lost | 1 | 1 | 1 | 4 | 1 |
| missense_variant | 2 | 2 | 2 | 5 | 1 |
| inframe_insertion | 2 | 2 | 1 | 5 | 1 |
| inframe_deletion | 2 | 2 | 1 | 5 | 1 |
| 5_prime_UTR_variant | 2 | 2 | 1 | 1 | 5 |
| 3_prime_UTR_variant | 2 | 2 | 1 | 1 | 5 |
| synonymous_variant | 2 | 2 | 2 | 2 | 5 |
| intron_variant | 2 | 2 | 2 | 2 | 5 |
| regulatory_region_variant | 2 | 2 | 1 | 1 | 5 |
| intergenic_variant | 1 | 1 | 1 | 1 | 5 |

**<u>Disease-associated variant consequences:</u>**

Once the variant classes associated with the disease are known, map these to the high-level terms using the matrix above.

High level terms to describe variant consequences:

| |
|---|
| • Altered gene product level<br>- Unspecified change in gene product level<br>- Decreased gene product level<br>- Absent gene product<br>- Increased gene product level<br>• Altered gene product sequence |
| • Functionally Normal |

Notes:

**Altered gene product level** - A sequence variant that alters the level or amount of gene product produced. This high-level term can be applied where the direction of level change (increased vs decreased gene product level) is unknown or not confirmed

e.g. promoter or enhancer variants, some splice variants

**Increased gene product level** - a variant that increases the level or amount of gene product produced

e.g. non-disruptive gene duplications, some promoter or enhancer variants

**Decreased gene product level** - a sequence variant that decreases the level or amount of gene product produced

e.g. a 5'UTR variant that reduced protein levels by disrupting translation, a 3'UTR variant that affects RNA stability, splice variants that decrease but do not stop expression, variants leading to nonsense mediated decay (NMD)-competent premature termination codon (PTCs), or gene-disrupting structural variants.

**Absent gene product** - a sequence variant that results in no gene product.

e.g. whole gene or other large scale disruptive structural variant, variants producing NMD-competent PTCs

**Altered gene product sequence -** a sequence variant that alters the sequence of a gene product.

e.g. missense variants, NMD-incompetent PTCs, and other length-changing variants (inframe indels, stop loss)

Downstream mechanisms are then diverse: functionally null - misfolded, mislocalized, inactive, hypomorphic; disruptive presence of abnormal protein (gain-of-function (GoF), dominant negative) etc.

**Functionally normal** - a sequence variant that is not expected to alter gene product sequence or levels

e.g. a synonymous variant

**Narrative summary of molecular mechanisms:**
Summary of mechanism for gene-disease pair. For example, 'Mechanism is likely loss of function of *NF1* due to reduction/absence of gene product or altered gene product sequence.'
Mention of specific mechanisms such as 'dominant negative' can be recorded here as well as any other useful information captured in the literature review section. Please record information on structural variants if they are relevant for this gene-disease pair.

**Record information about other ACMG evidence types if it is available (this will not be available for all gene-disease pairs).**

Examples and notes:

**For PM1:**
- Is there a mutational hot spot for this gene-disease pair? If so, is there a guideline and genomic coordinates?
- Is there sufficient data to calculate etiological fraction by domain? If so please record it here

Notes:
Etiological fraction applied per domain/region can provide following levels of evidence for PM1: Strong >0.95, moderate >0.90, supporting >0.80.
For more information on Etiological Fraction see: https://genomemedicine.biomedcentral.com/articles/10.1186/s13073-019-0616-z (PMID: 30696458)
See Walsh et al 2020 PMID: 32893267 for specific examples of this for LQTS and Brugada genes, in particular table 2: https://www.nature.com/articles/s41436-020-00946-5/tables/2

**For PVS1:**
- Is there any important transcript information, i.e. are there certain transcripts that are or are not relevant to this disease.

Notes:
For example in the *TTN* gene, truncating variants are only known to cause disease if located in exons that are constitutively expressed in the heart (i.e. PVS1 doesn't always apply)
See Whiffin et al 2018 PMID: 29369293 for examples of tailored application of PSV1

**For BS1/PM2**:
- What is the maximum credible population allele frequency for this disease?

Notes:
maximum credible population AF = prevalence x maximum allele contribution x 1/penetrance
See https://www.nature.com/articles/gim201726 (PMID: 28518168) for more information about maximum credible population AF and examples
See Walsh et al 2020 PMID: 32893267 for an example of filtering allele freq thresholds for LQTS and Brugada genes.

**For PP2**:

- If there are no mutational hotspots, what is the etiological fraction for the whole gene (this may not be available for very rare conditions where there may be insufficient case numbers to do the analysis).

Notes:
Etiological fraction applied per gene can provide following levels of evidence for PP2:
Strong >0.95, moderate >0.90, supporting >0.80.
For more information https://www.ncbi.nlm.nih.gov/pmc/articles/PMC5116235/ (PMID: 27532257)

**General notes:**
-If monoallelic and biallelic inheritance can cause the same disease, they should be recorded as separate entities if biallelic variants lead to a different phenotype (not just a change in severity)
-If a dominant variant can also be seen on both alleles but the outcome is essentially the same disease, then this should be categorised as one entity using dominant and monoallelic.

For example:
AD and AR *DSC2* causing isolated ARVC are one disease gene pair
AR *DSC2* causing ARVC with cutaneous manifestations is a separate disease gene pair.
